# Unravelling Causal Associations between Population Mobility and COVID-19 Cases in Spain: a Transfer Entropy Analysis

**DOI:** 10.1101/2023.10.09.23296732

**Authors:** Miguel Ponce-de-Leon, Camila Pontes, Alex Arenas, Alfonso Valencia

## Abstract

Human mobility is a well-known factor in the spread of infectious diseases. During the COVID-19 pandemic, the rapid spread of the SARS-CoV-2 virus led to healthcare systems collapsing in numerous countries, such as Spain and Italy, resulting in a significant number of deaths. To avoid such disastrous outcomes in the future, it is vital to understand how population mobility is linked to the spread of infectious diseases. To assess that, we applied an information theoretic approach called transfer entropy (TE) to measure the influence of the number of infected people travelling between two localities on the future number of infected people in the destination. We first validated our approach using simulated data from a SIR epidemiological model and found that the mobility-based TE was effective in filtering out non-causal influences that could otherwise arise, thereby successfully recovering the epidemic’s spreading patterns and the mobility network topology. We then applied the mobility-based TE to analyse the COVID-19 pandemic in Spain. We identified which regions acted as the main drivers of the pandemic at different periods, both globally and locally. Our results unravelled significant epidemiological events such as the outbreak in Lleida during the Summer of 2020, caused by the influx of temporary workers. We also analysed the effects of a non-pharmaceutical intervention in Catalunya, using mobility-based TE to compare the infection dynamics with a control region. These results help clarify how human mobility influences the dynamic spread of infectious diseases and could be used to inform future non-pharmaceutical interventions.

## Introduction

The COVID-19 pandemic emerged as an unprecedented global health crisis, challenging healthcare systems, economies, and societal norms worldwide. As countries strive to mitigate the impact of future epidemics, integrating innovative data-driven strategies becomes imperative (1). One promising approach is the use of anonymized phone-based mobility data (APMD) in conjunction with non-pharmaceutical interventions (NPIs) to inform and optimize public health responses (2–5). Over the last decade, applications of APMD have emerged as a tool to develop models of human mobility (6, 7), to analyze the spatial structure of cities (7, 8), to infer friendship networks (9), mapping dynamic population (10), to predict poverty and wealth (11) and in disaster management (12). In epidemiology, this kind of data has been integrated into different classes of epidemiological models to account for population mobility between regions (13–18).

While it is clear that population mobility plays a crucial role in the spreading process of infectious diseases, inferring a causal influence or coupling between population mobility patterns and regional disease outbreaks is far from trivial. Nonetheless, finding a way to detect whether an increase in the number of cases in a region is likely to ‘cause’ an outbreak in a different region is critical to help in the development of effective NPIs (3, 19). We found that the big volumes of data in the form of time series generated during the COVID-19 pandemic (20–24) constitute an excellent opportunity to test and develop methods that can inform about possible causal connections between the different observables such as population mobility, novel variants, regional outbreaks, and so on.

Causality inference is a long-standing problem with roots in different disciplines including philosophy, economics, statistics, and biology (25). There are several ways to define causality between two variables *X* and *Y* (26). A widely adopted definition states that *X* causes *Y* if an intervention on *X* produces a change in *Y* (25). Based on this definition, causal relations can be inferred from time series data using statistical and data-driven approaches. Since these approaches do not rely on any explicit mechanistic model of the processes generating the data, they are commonly referred to as ‘model-free’ methods (see [26, 27] for an extensive review). Model-free methods can be broadly grouped into two classes (28).

The first class includes methods that rely on specific assumptions about the processes generating the observed data. One example is Granger Causality (GC), a statistical hypothesis test of whether *X* improves the forecasting of *Y* (29, 30). Even though GC has been successfully applied in many different domains, it relies on several restrictive assumptions, including linearity, and stationarity among others (see [28] for an extensive review). Furthermore, GC assumes that some properties of *X* can change independently of *Y* and that the flow of information is essentially unidirectional. In complex dynamical systems, however, the interactions between the system’s components can be bidirectional (leading to cyclic flows of information) and change over time (31, 32). Moreover, dependencies between the system components are typically nonlinear and therefore most of the GC’s assumptions do not hold. In the case of epidemic processes, nonlinear dependencies and transient coupling between regions are expected due to the complex interplay between daily population mobility and social interactions that lead to the spreading process. Therefore, classical methods for causal inference such as GC become problematic in complex systems where cause and effect can be entangled (33).

The second class include information-based methods that quantify the extra information about the dynamics of *Y* provided by the knowledge of *X* (26, 34). One example is Transfer Entropy (TE), an approach for measuring the amount of directed (time-asymmetric) transfer of information between two random processes (35). TE has been proven to be a generalization of GC and, for Gaussian variables, both approaches are entirely equivalent (36). However, TE has the advantage that it does not have any underlying assumptions on the nature of the processes generating the observed data (37). As a consequence, TE has been applied to study causal relations in non-linear systems in different fields including neuroscience (34), complex systems (38, 39), economics (40), among others. More recently, TE has been also applied in the context of epidemic modelling, to study the age structure transmission during the autumn wave of the 2009 influenza pandemic in the USA (41).

Inspired by previous work on the use of TE to measure the temporal and structural signatures of collective social events (42), we applied TE to measure how population mobility drives the spread of an epidemic process. Specifically, we the TE to measure the information flow between the number of infected individuals travelling between two regions, and changes in the rate of new infections at the destination. We first tested our approach in simulated scenarios using a meta- population epidemiological model. We found that integrating the structure of the population mobility network is essential for discarding non-causal relations between epidemiological outbreaks in different locations.

We then applied our approach to study the COVID-19 pandemic in Spain using real data including time-series of COVID-19 daily cases and origin-destination daily mobility matrices reconstructed from APMD (24). We characterize different information transfer patterns that arise between neighbour and non-neighbour regions together with their implications in the spreading dynamics. We used our approach to detect which provinces acted as main drivers of the different COVID-19 outbreaks in Spain and found that the results are in agreement with many known epidemiological events. Finally, we applied our approach to analyse the consequences of an NPI applied in Catalunya (4). We observed that the NPI caused a temporary decoupling between movement and disease spreading patterns, and identified which regions changed their status from drivers to neutral after the application of NPI. Altogether, we showed that the mobility-based TE approach allows inferring causal relationships be tween population mobility and the spread of infectious diseases and thus it can be used as a tool to support decision-making and NPI design in future epidemics.

## Results

### Recovering disease spreading patterns with mobility-based transfer entropy

Given a set *P* of interconnected subpopulations or regions, we use *I*_*y*_(*t*) to denote the time series of daily new cases of COVID-19 reported at region *y ∈ L* (Fig. 1A), and *M*_*x,y*_(*t*) for denoting the number of daily trips from *x* to *y* at date *t*. We reasoned that a region *x* will only have a coupling effect on the number of cases in the region *y* if and only if there are infected individuals in *x* and also if there are persons travelling from *x* to *y*. We then use the number of reported cases over the last ten days *I*_*x*_ (*t*) in region *x* and the trips *M*_*x,y*_(*t*) to calculate the mobility-associated risk *R*_*x,y*_(*t*) between each pair of regions (Fig. 1B), as previously defined (see Materials and Methods and (24)). This quantity estimates the potential number of infected individuals travelling from source *x* to target *y* on the date *t*. Then, to evaluate whether a given region *x* has influenced the dynamics of new infections observed in *y*, we measure the *TE*_*x,y*_(*t*) between *R*_*x,y*_(*t*), *i*.*e*. the mobility-associated risk from *x* to *y*, and *I*_*y*_(*t*) (Fig. 1C). We choose *R*_*x,y*_(*t*) as the time series on the source because this quantity integrates information on the number of cases in the source region *x* together with a structural connection to the target region *y* given by the number of trips.

**Fig. 1.**
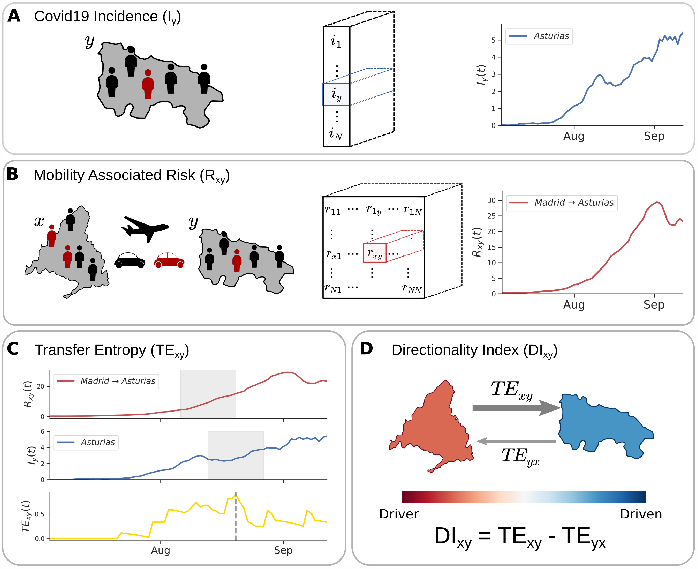
Mobility-based Transfer Entropy for studying epidemic process using real data on cases and mobility. Panel A shows the incidence of COVID-19 ((*I*_*y*_)) for each region *y*, expressed as cases reported per 100,000 inhabitants. Panel B shows the mobility-associated risk (*R*_*x,y*_), which is calculated for each pair of regions (*x, y*) over time taking into account the proportion of infected people travelling from *x* to *y*. Panel C shows the Transfer Entropy *TE*_*x,y*_, which measures the mobility-related influence *x* has over *y. TE*_*x,y*_ values are high when past values of *R*_*x,y*_ help reduce the uncertainty about future values of *I*_*y*_. Lastly, panel D shows the Directionality Index *DI*_*x,y*_, which measures the asymmetry in entropy transfer between a pair of patches. Low values of *DI*_*x,y*_ characterise *x* as a driver of infection in *y*, while high values of *DI*_*x,y*_ mean infections in *x* are being driven by *y*.

Since the spreading of an epidemic between a set of interconnected regions is a dynamic process, we expect the influence between regions to change as the epidemic evolves and thus, we measured the *TE*_*x,y*_ using a sliding window of size *ω* (Fig. 1C). To account for the delay between any changes in the daily number of cases reported in a region *y* due to the arrival of the infected individual from a region *x* we introduce a time delay *δ*. Then, for a source region *x* and a target region *y* we measured the *TE* considering the values of *R*_*x,y*_ in the time interval [*t, t* + *ω*] and the number of cases *I*_*y*_ in the time interval [*t* + *δ, t* + *ω* + *δ*] (Fig. 1C). Therefore, for each window, we measure the effect of the trips from *x* to *y* in the rate of new infections observed in *y* after a delay of *δ* days. Since the influence between two regions (*x, y*) can be bidirectional, we calculate the asymmetry in the transfer of entropy using the directionality index *DI*_*x,y*_(*t*) (see equation 2) which measures the directional flow of information, i.e. it gives information about which region is influencing (driving) or being influenced (driven) by the other one (Fig. 1D). Finally, since the *DI*_*x,y*_(*t*) matrix for a given date is symmetric in the absolute value (i.e. *DI*_*x,y*_ = *−DI*_*y,x*_) we consider only the positive entries 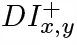 (see equation 3) to quantify the net directional flow of information between a pair of regions.

To evaluate the performance of the mobility-based TE as a measure of causal influence between population mobility and the spreading of an epidemic, we first applied our approach to the controlled case of a simulated epidemic process on a meta-population model (Fig. S1). Using the simulated epidemic we measured the *TE* between a source region *x* and a target region *y* in two different ways. First, we measure the TE using the time series of incidences *I*_*x*_(*t*) and *I*_*y*_(*t*) without considering the mobility, and then between the mobility-associated risk *R*_*x,y*_(*t*) from the source to the target and the incidence in the target region *I*_*y*_(*t*). We observed that while the TE measured between the time series of the incidence of the simulated disease in different locations gave rise to many indirect influences, the mobility-based TE, as proposed here, correctly recovers the spreading patterns of the epidemic process, as well as, the structure of the mobility network (Figs. S2, S3 and S4). Further details about all different simulated scenarios can be found in the Supplementary Information. Altogether, this analysis showed that the mobility-based TE is effective in recovering the spreading patterns between different regions in a simulated epidemic process.

### Information flows between neighbour and non-neighbour regions in Spain during the COVID-19 pandemic

Here, we used the mobility-based TE to investigate the role of population mobility in the spread of SARS-CoV-2 in Spain during the first waves of the COVID-19 pandemic. We used phone-based anonymized mobility data to estimate the daily number of travels between regions together with a time series of daily reported cases for the different regions (see Material and Methods Sections for more details). We first focus the analysis at the country level considering as regions the fifty provinces of Spain and excluding the autonomous cities of Ceuta and Melilla (see Supplementary Fig. S5 and Material and Methods Sections). We measured the *TE*_*y,x*_ between the mobility-associated risk *R*_*x,y*_(*t*) and daily reported COVID- 19 cases *I*_*y*_(*t*) between all pairs of provinces in Spain using a sliding window of *ω* = 14 days and a delay *δ* = 7 days (see Materials and Methods for further details on the calculation). We found that the patterns of entropy transfer are robust to changes in the values of *ω* and *δ*, which cause *TE* peaks to slightly slide to the right or to the left (see Supplementary Figure S6).

We first compared the dynamics between mobility, cases and TE between neighbouring and distant regions. The results show that neighbouring regions, e.g. Madrid and Guadalajara (for a map of all the provinces of Spain see Fig. S5) exhibit similar flows of people in both directions characterized by high values (Fig. 2A, right panel). This pattern is consistent during the whole period of study and is mainly driven by daily commuting. Closer regions also exhibit similar trends in the normalized number of daily reported cases, in contrast with distant regions where the waves of cases occurred in different time periods (Fig. 2B right and left panel). Non-neighbour regions, on the other hand, show different mobility patterns between each other and also exhibit a lower number of trips than neighbour regions (Fig. 2A, left panel). The time series of mobility-associated risk *R*_*x,y*_(*t*) exhibits similar patterns to those observed in the daily mobility and daily reported cases (Fig. 2C). Similar patterns are observed when comparing Barcelona (Supplementary Fig. S7), Valencia (Supplementary Fig. S8) and Granada (Supplementary Fig. S9) to neighbour and non-neighbour regions.

When we compared the information flow transferred between neighbour and non-neighbour regions we found that neighbouring regions transfer similar values of *TE*_*x,y*_ in both directions during most of the time. This symmetric behaviour results in values of the directionality index *DI*_*x,y*_(*t*) close to zero during long periods, or values that oscillate changing the direction of the coupling (Fig. 2D, left panel). We reasoned that neighbour regions have a strong bidirectional coupling due to daily population mobility and thus are close to a certain ‘equilibrium’ characterized by a *DI*_*x,y*_(*t*) close to zero.

Nonetheless, this symmetry breaks for specific periods, and one of the regions becomes a driver over its neighbour. For instance, during August and October, we observed peaks of *DI*_*x,y*_(*t*) from Madrid to Guadalajara (Fig. 2D, left panel). On the other hand, distant regions do not show bidirectional coupling and there is no information flow *TE*_*x,y*_ transferred in any direction during long periods. However, whenever we measure *TE*_*x,y*_ it is usually transferred in one direction indicating a directional coupling. For instance, if we compare Madrid and Asturias the results show that there is almost no transfer of entropy in any direction until the beginning of the summer (mid-July). During the summer the result shows a peak in the mobility associate risk from Madrid to Asturias driven by two factors: an outbreak of cases in Madrid together with a high increase in the number of trips between the two regions (Fig. 2A-C, right panels). The results also show that during this period the 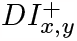 from Madrid to Asturias exhibits a peak (Fig. 2D, right panel), suggesting that the summer vacation trips could be related to the later outbreak of cases observed in Asturias. Therefore, while Madrid may have contributed to driving the increase of cases in Asturias during summer, Asturias did not influence Madrid. Further examples comparing neighbour and non-neighbour regions are provided in Supplementary Figures S7, S8 and S9. In general, we found that an asymmetry in the transfer of entropy between two regions could be an indicator that the source region may be driving, at least partially, changes in the rate of new infections in the target region.

**Fig. 2.**
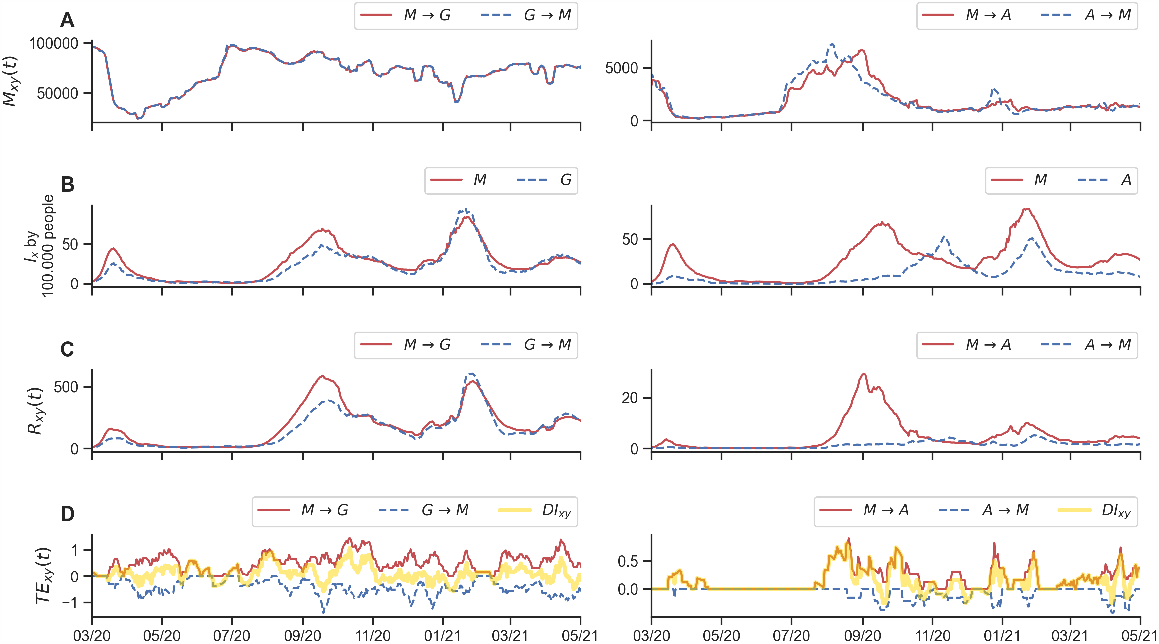
Time series of mobility, cases, and *TE* between neighbour and non-neighbour provinces. The figure depicts different time series between a pair of neighbour provinces Madrid-Guadalajara (right panels), and non-neighbour regions Madrid-Asturias (left panels). From top to bottom, the figure depicts the number of daily trips (A), normalized new cases (B), the associated risk between (C) and the transfer of entropy *TE*_*x,y*_ (D). For visualization purposes, *TE*_*x,y*_ is represented as positive values whereas *TE*_*x,y*_ (the opposite direction) is represented as negative ones. The yellow line in D represents the directionality index *DI*_*x,y*_. In the legends, the labels M, G and A, correspond to Madrid, Guadalajara and Asturias respectively.

### Detecting outbreaks and its main drivers during the COVID-19 pandemic in Spain

We used our approach to analyze the spreading patterns of the COVID-19 pandemic in Spain during the first three waves. Specifically, we focus on detecting which provinces drive the spread of cases and which ones were driven during the different phases of the studied period. We first calculated the directionality index *DI*_*x,y*_ between all the provinces as a way to measure the coupling between the different regions. Then, to evaluate the role of each province we aggregated the total *DI*_*x*_ for each province on each day, and used these values to identify which provinces were drivers (*DI*_*x*_ *>* 0) and which ones were being driven (*DI*_*x*_ *<* 0) during the different phases of the pandemic. Figure 3A shows the *DI*_*x*_ profile for each province, ordered in decreasing order by the total *DI*_*x*_ aggregated across time, and thus main drivers are placed on the top and provinces that were mainly driven are placed at the bottom (Fig. 3A).

**Fig. 3.**
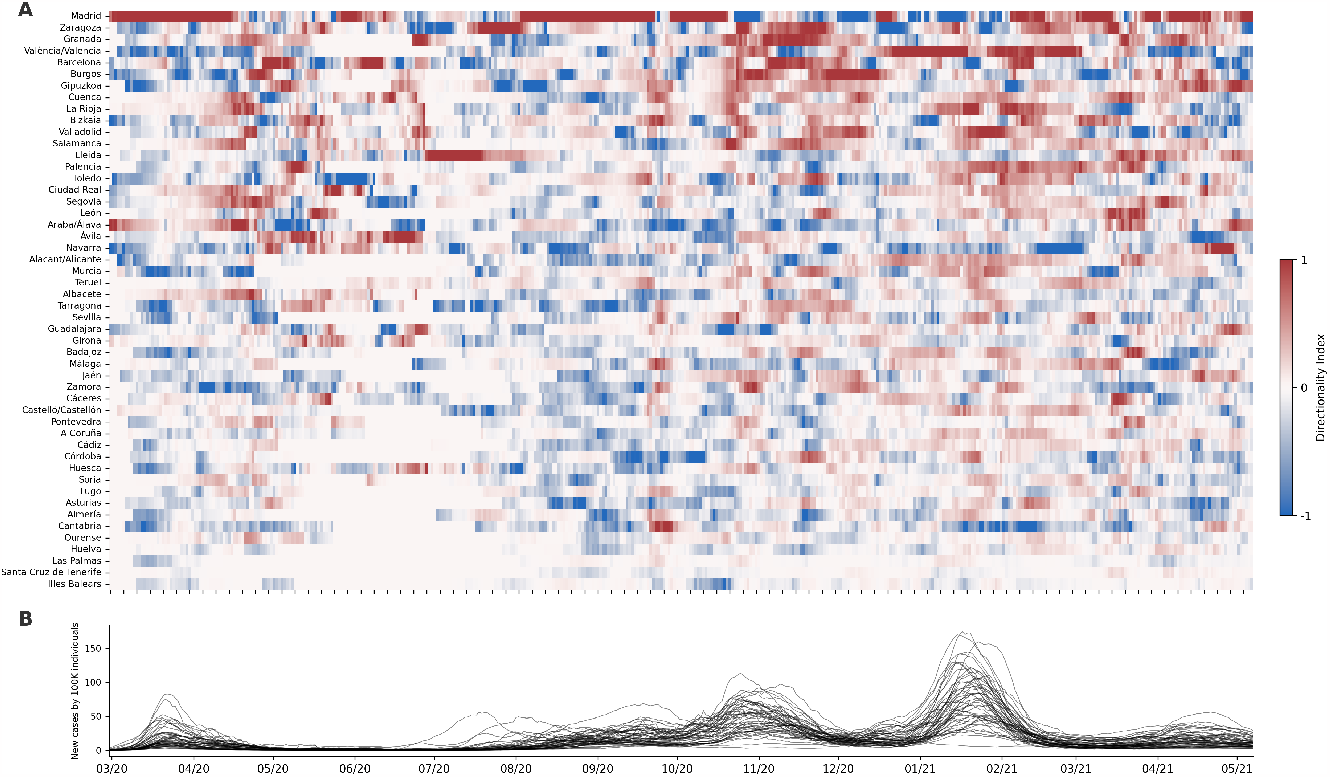
Dynamics of the COVID-19 pandemic in Spain. Panel A shows the average normalised directionality index *DI*_*x*_ for each province during the COVID-19 pandemic calculated. Provinces are ordered by total *DI*_*x*_. *TE* was calculated using parameters *δ* = 7 days and *ω* = 14 days. Panel B shows the average number of daily COVID-19 by 100k inhabitants for the same period.

The results show that Madrid was the major driver of cases throughout the pandemic followed by Zaragoza, Granada, Valencia and Barcelona (Fig. 3A). Madrid, Barcelona and Valencia are the top three most populated provinces in Spain and therefore are expected to be at the top because their population have more chances of “exporting” infected individuals to other provinces. On the other extreme, we found the Balearic and the Canary Islands to be the most driven regions of Spain together with distant distant provinces such as Huelva and Ourense. This is also an expected result since because of their isolation, the islands are not likely to be driver regions. It is worth noting that among the top five driver provinces, we also found Zaragoza and Granada which are mid-size provinces. This suggests that the population size is not the only factor to become a driver. Granada is one of the most tourist cities in Spain which may explain its role as one of the main drivers. In the case of Zaragoza, it is a province that connects Barcelona and Madrid and, as will be discussed later in this section, it played a critical role as a driver in the second wave. The results also show that although Madrid was the main driver, it played different roles during the second and third waves of the pandemic (Fig. 3A and B). During the second wave, it acts as a strong driver while during the third wave, it is mainly driven. These results agree with the population dynamics of Spain where people usually move out from Madrid during Summer Vacations and travel to Madrid during Christmas.

When we focus the analysis on the beginning of the pandemic the results show that, together with Madrid, the provinces of Álava and La Rioja are also found to be among the main drivers. Interestingly, this is in good agreement with the chronology of the pandemic in Spain (43). According to a press article published in El Pais newspaper, at the end of February 2020, a funeral held in Vitoria (the capital of Álaba) was identified as the largest episode of the spread of the coronavirus recorded in the country up to that time (44). In this event, more than 60 attendees at the ceremony were infected by the pathogen, 38 of whom live in Haro and Casalarreina (La Rioja) and another 25 in Álava.

Given the likely influence of population movement at the onset of the second COVID-19 wave in Spain, we decided to analyse this period in greater detail. At the beginning of Summer, our results show that Lleida started acting as an important driver, probably driving an outbreak of cases in Zaragoza and later in Madrid. This event is known to have happened due to the arrival of seasonal workers who went to Lleida and Huesca and then got back to Zaragoza and Madrid (45). Actually, in Figure 3B the peak of cases observed in July corresponds to Lleida (46). The spreading pattern of this event can be seen in Figure 4, which shows all Spanish provinces coloured by daily COVID-19 incidence and the significant net flow of information 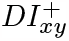 between them. To corroborate the spreading patterns shown in Figure 4, we looked for the main three sources of entropy for Zaragoza during June and July and found that they correspond to Lleida, Huesca and Barcelona (Fig. S10). The same analysis performed for Madrid shows that Zaragoza, Toledo and Barcelona were the main sources of entropy during July and August (Fig. S11). Altogether, our results showed how the mobility-based TE can be used to uncover the spatiotemporal patterns of the spreading of an epidemic process such as the COVID-19 pandemic.

**Fig. 4.**
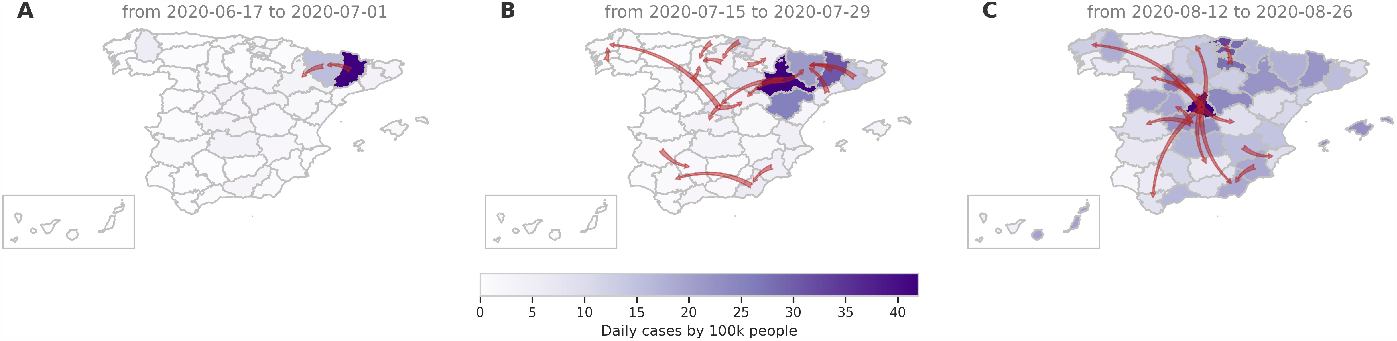
Transfer of entropy between Spanish provinces during the Summer. The maps show Spanish provinces coloured by the number of daily COVID-19 cases by 100k inhabitants. Red arrows indicate the net flow of information 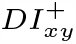 between provinces during a period of 14 days starting at different dates. Only 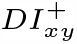 values above the 0.95 percentile are represented in each map.

### Evaluating the impact of non-pharmaceutical interventions: the case of closing bars in Catalunya

Between October 16 and November 25, 2020, the local government of Catalunya applied the policy of closing all bars and restaurants with the aim of flattening an outbreak of COVID-19 cases. In previous work, we analyzed the effect of the policy on population mobility and found that the movements were reduced during this period, unlike Madrid, the control case, where no policy was applied (4). Herein, we applied the mobility-based TE to study the effect of the policy. We calculated the TE between the Basic Health Areas (BHA) for the region of Catalunya and Madrid using population mobility and COVID-19 cases, reported at a higher spatial resolution (see Materials and Methods). Figure 5 shows the number of trips *M* (*t*) per inhabitant, the daily incidence, the mobility-associated risk *R*(*t*), and the average *DI*^+^, for Catalunya and Madrid. The results show that in Catalunya the average mobility *M* (*t*) starts decreasing at the beginning of the policy whereas in Madrid, where no policy was applied, the mobility increased during this period (see Fig. 5A and Supplementary Fig. S12A for a wider period). The data also showed that during the two first weeks of the policy, the daily incidence was growing in Catalunya, while in Madrid the daily cases were already decreasing when the policy was applied (Fig. 5B). In fact, the second wave of COVID-19 observed in Madrid started in August and reached its maximum at the end of September (see Supplementary Fig. S12B). The mobility-associated risk *R*(*t*) shows similar trends that those described for the incidence (Fig. 5C).

**Fig. 5.**
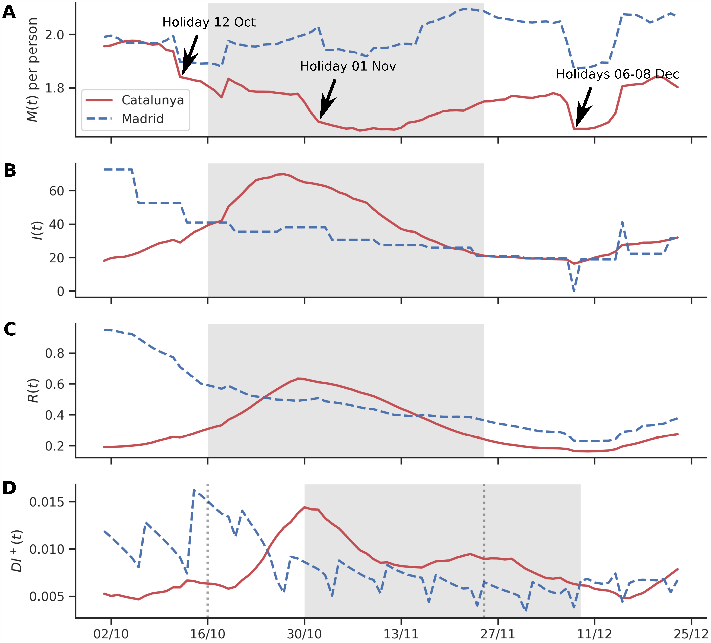
Impact of the temporary closing of bars and restaurants in Catalonia. Panel A, B, C and D show the total net flow of information *DI*^+^(*t*), the number of trips M, the mobility associated risk R and the number of new cases by 100k inhabitants I across time, respectively, for all basic health areas in Catalonia (in red) and Madrid (in blue) during the period of the closing of bars in Catalonia (a policy applied between October 16 and November 25, 2020).

Two weeks after the application of the policy in Catalunya the daily incidence *I*(*t*) and the mobility-associated risk *R*(*t*) for this region changed their trends and started decreasing. We also calculated the rate of change for the daily incidence and found that a week after the policy was applied, there was an abrupt deceleration in the rate of change of new cases in Catalunya whereas in Madrid there were no changes (see Supplementary Fig. S12C). We calculate the correlation between mobility and the rate of change for the daily incidence, for the whole of 2020 and for the period when the policy was applied in Catalunya (Supplementary Fig. S13C). The results showed a moderate correlation when the whole year was considered with *R* = 0.25 and *R* = 0.14 for Catalunya and Madrid, respectively. Strikingly, when the correlation is calculated throughout the policy we found a very strong correlation between mobility and the rate of change for the daily incidence with an *R* = 0.84 for Catalunya (*p* = 5*e* − 11), while for Madrid the correlation was close to zero and statistically not significant.

When we analyzed the net flow of information between the BHAs, we found that the average *DI*^+^ started decreasing in Catalunya 14 days after the application of the policy (Figure 5D). This is an expected result since the TE was measured using a sliding window of *ω* = 14 days. In the case of Madrid, we also observed a decrease in the average *DI*^+^, probably driven by the decrease in daily cases. However, when we compared the average *DI*^+^ transferred during the first two weeks of the policy with the average *DI*^+^ transferred during the last two weeks of this period, for Catalunya (see Supplementary Fig. S14), we found a significant decrease for Catalunya (*p* = 2.3*e* − 22). Madrid also showed a statistically significant *DI*^+^ reduction for the same period (*p* = 4.7*e* − 03), but the magnitude of the reduction is lower than that observed in Catalunya.

Taking a closer look at which BHA changed their driver/driven patterns during this period, it is possible to see that BHA located in municipalities which are on the outskirts of the city of Barcelona changed their status of drivers to neutral. Some examples include Sant Joan Despí, Badalona and Matorell (Fig. S15). This is probably because people that live in these locations stopped going to the centre of Barcelona to eat out and drink. These results show that the dynamics of the epidemic in Catalunya have been affected by the temporary closing of bars and restaurants. There is a reduction in the total number of trips in Catalunya following the application of this measure which does not seem to be explained by other known factors. We hypothesise that this reduction in population movement causes a decoupling between the movement patterns and the incidence patterns which helps revert the rising trend in the number of cases. All of this is reflected in the changes observed in the *DI*^+^ patterns.

## Conclusions

In this work, we introduced mobility-based TE to investigate potential causal relationships between population mobility and the spreading of infectious diseases. We tested the proposed methodology on simulations of an epidemic process using meta-population models considering different topologies of the mobility network. Our results showed that, if the TE is measured between the incidence time series without considering mobility, the results include a high number of spurious correlations pointing to non-causal relationships. Therefore, we conclude that for detecting possible causal relationships, or couplings, between regions during an epidemic process, the structure of the mobility network must be considered explicitly.

When applied to the COVID-19 pandemic dataset, our approach allowed us to identify the regions that acted as the main drivers during different periods of the pandemic in Spain. Additionally, we investigated the effect of the policy of closing bars and restaurants in Catalunya. Our results showed that reductions in mobility were followed by a strong decrease in the information transferred and a reduction in the daily incidences when compared to a control region. Altogether, we found that the net transfer of information between a source and a target region may be an indicator that the infected population coming from the source region could be driving, at least partially, changes in the infection dynamics in the target region.

It is worth reiterating that identifying causal relationships is a big challenge, especially in the case case of complex, dynamical systems containing real world data. We acknowledge that our approach has some limitations and, therefore, the results obtained should be carefully analyzed before drawing any conclusions. First, one must be aware that the signal captured by the TE does not necessarily imply causation. We partially mitigate this problem by removing non-causal associations that would otherwise arise if we did not take mobility patterns into account in the analysis. Second, the interpretation of the signal extracted from the TE analysis is not straightforward. The value of the TE will rise whenever changes in movement patterns influence changes in infection patterns. This influence, however, can be of different types. During an outbreak, a reduction in movement might cause a reduction in the number of cases, but right before an outbreak, an increase in movement might lead to the initial rise in cases, just to give a couple of different examples.

Despite these limitations, the patterns extracted through our analysis are robust enough to correctly unravel the dynamics of known epidemiological events and, therefore, should be considered as possible foundation of a framework to inform future non-pharmaceutical interventions and strategies aimed at curtailing infectious diseases transmission.

## Material and Methods

### Transfer Entropy

Transfer entropy (*TE*) is an information-based method that measures the directional flow of information between two random time-dependent processes *X*(*t*) and *Y* (*t*) (35). More precisely, the *TE* measures the amount of uncertainty reduced in future values of *Y* when considering the past values of *Y* and the past values of *X*. Therefore, it can provide information about possible causal connections between the different observable variables. The *TE* is calculated by computing the joint and conditional probabilities of the sequence indices from the relative frequency of symbols in each sequence:

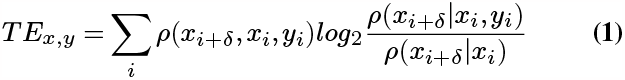

An essential property of Transfer Entropy is its asymmetry, i.e. *TE*_*x,y*_ ≠ *TE*_*y,x*_, this property allows quantifying the directional coupling between components of complex systems. The directionality index, *DI*_*x,y*_, measures the asymmetry in the transfer of information between two processes and is given by:

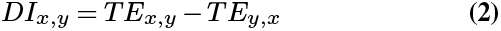

A positive value of *DI*_*x,y*_ means process *X* is transferring a net flow of information to process *Y*, whereas a negative value of *DI*_*x,y*_ means that the net flow of information goes in the from *Y* to *X*. Therefore, we use the net directional flow of information, 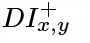, to measure the dominant direction of the information flow between two components *X* and *Y*, given as follows

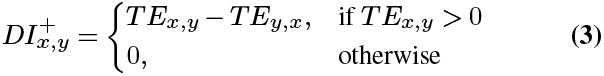

In this work, we applied a mobility-based TE approach to investigate the relationship between population movement patterns and the spreading of COVID-19 cases in Spain. Specifically, to evaluate the effect of mobility in the spreading dynamics of COVID-19 between geographic zones in Spain throughout the pandemic, we calculated the TE between the mobility-associated risk *R*_*x,y*_ (*t*), as given by Eq. 1, and incidence *I*_*y*_ (*t*), for each pair of zones *X*, and *Y*.

The intuition is that, in the target region, *Y*, an outbreak may be caused by infected individuals coming from one (or several) source region *X*. In such scenarios, we expect to measure a transfer of entropy *TE*_*x,y*_ between time-series *R*_*x,y*_ (*t*) and *I*_*y*_ (*t*) for some of the source region *X*. The calculation of the TE requires the domain of the input signals to be a discrete alphabet and, therefore, before applying the TE, the time series should be discretized.

Finally, since the spreading of an epidemic between a set of interconnected subpopulations is a dynamic process, we expect the influence between subpopulations to change as the epidemic evolves. Therefore, we use a sliding window of size ω and measured the *TE*_*x,y*_ considering the values of *R*_*x,y*_ (*t*) in the time interval [*t, t* + *ω*] and the evolution in the number of cases after a time delay *δ, i*.*e*. in the time interval [*t* + *δ, t* + *δ* + *ω*] (see Fig. 1C). For all analyses, we considered parameters *ω* = 14 days and *δ* = 7 days. A time delay of 7 days was chosen based on the median infectious period for asymptomatic COVID-19 cases which is estimated to be between 6.5 and 9.5 days (47). A sliding window size of 14 days was chosen as a minimum interval to avoid artefacts (since for some regions the cases were reported weekly) and still have a fine grain entropy transfer estimation across time.

### Datasets

The COVID-19 Flow-Maps is a cross-referenced geographical information system which includes time-series accounting for population mobility and daily reports of COVID-19 cases in Spain at different scales of time spatial resolution (24). In this work, we have retrieved and used different data sets from COVID- 19 Flow-Maps available at Zenodo. The datasets include daily reports on COVID-19 cases (48), population (49) and population mobility (50) at different levels of geographic resolution (51). Since the different data records span across different time periods, we selected the maximum possible time interval that includes reported entries in all the datasets, including, case reports, population and mobility. We trimmed all the data records to span from 01/03/2020 to 01/05/2021, the maximum time range to which mobility and population data sets are available. Table 1 summarized the different datasets used in this work which are explained in the further subsections.

**Table 1.** Summary of the different datasets used in this work.

| Region | Dataset | Layer | N° patches |
| --- | --- | --- | --- |
| Spain | COVID-19 cases | Provinces | 50 |
| Spain | Population | Provinces | 50 |
| Spain | Mobility | Provinces | 50<sup>†</sup> |
| Catalunya | COVID-19 cases | BHA | 374 |
| Catalunya | Population | BHA | 374 |
| Catalunya | Mobility | BHA | 374 |
| Madrid | COVID-19 cases | BHA | 286 |
| Madrid | Population | BHA | 286 |
| Madrid | Mobility | BHA | 286 |

<sup>†</sup> The cities of Ceuta and Melilla were excluded from the study because of the low coverage founded in the phone-based mobility data.

#### COVID-19 Case reports

We retrieved daily cases for all of Spain reported at the level of provinces and at the level of Basic Health Areas (BHA) for Catalunya and Madrid (48). Each record corresponds to a geo-referenced time series where each record has an associated date, the corresponding identifier of the layer, a code of the zone (*i*.*e*. a province or BHA) and the number of cases reported on that date (daily incidence). To avoid possible artefacts in the cases reports, due for instance to delay in reporting, we applied a rolling average using a 7-day sliding window. In this way we smoothed the time series, removing outliers that might impact the analysis. Rolling averages were calculated using standard utilities from the Pandas Python package (see **Software and computational resource** subsection). Through the text, we will refer to the daily number of cases reported for the region or patch *x* ∈ *L* in date *t* ∈ *T* as *I*_*x*_(*t*), and as 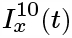 to the accumulated incidence over the ten days that preceded *t*.

#### Population and Daily Mobility Data

Mobility and population data records come from a study conducted by the Spanish Ministry of Transport, Mobility and Urban Agenda (MITMA, *Ministerio de Transportes, Movilidad y Agenda Urbana*) which analyses the mobility and distribution of the population in Spain from February 14^th^ 2020 to May 9^th^ 2021 (https://www.mitma.gob.es/ministerio/covid-19/evolucion-movilidad-big-data). The data records are based on a sample of more than 13 million anonymized mobile-phone lines provided by a single mobile operator whose subscribers are evenly distributed and include two different mobility indicators. Daily origin-destination matrices (ODM) account for the number of trips between 2850 *mobility areas* that cover almost the entire territory of Spain, reconstructed by combining cell phone antenna coverage areas with districts and municipalities. The second mobility indicator reports for each *mobility areas* the total number of persons that have performed 0, 1, 2 or more than 2 trips in a given date. The indicator accounts for the fractions of people performing at least one trip or none, as well as the estimated total population in that zone for the given date. In this work, we used the mobility indicators (50) and population data (49), projected into provinces and BHA. With respect to mobility indicators and population, we will use the following notations: *M*_*x,y*_ (*t*) is the daily number of trips from zone *x* to *y* with both *x, y ∈ L* and reported at date *t*, and will use *P*_*x*_ to refer to the total population for zone *x ∈ P*.

### Mobility-associated risk

To assess the effect of population mobility on the spreading and outbreaks of COVID-19, we have previously introduced the Mobility Associated Risk (MAR) (24). The MAR *R*_*xy*_ (*t*) is an estimator of the potential number of infected individuals moving from zone *x* to *y* on the date *t* and is given by the following expression:

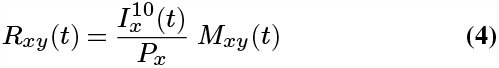

where the first term represents the density of cases in the source zone *x* (i.e. the number of active cases over the last 10 days 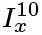 per the total number of inhabitants *P*_*x*_) and *M*_*xy*_ (*t*) is the number of trips from *x* to *y*. Finally, for a given date *t*, the values *R*_*xy*_ (*t*) can be arranged in a |*L*| × |*L*| matrix *R*(*t*) which represents a directed weighted network where the nodes correspond to the different zones *x, y* ∈ *L* and the flow *R*_*xy*_ (*t*) between any pair of nodes (*x, y*) is the estimated number of infected subjects moving from the source *x* to target *y* at time *t*.

### Software and computational resources

Transfer entropy calculations were conducted using the PyInform python package, which is a wrapper for *Inform* is a C library that implements several information-based approaches for the analysis of complex systems (52). Data processing, analysis and plots were using the following Python packages: Geopandas, xarray, scipy, sklearn, matplotlib and seaborn. All the calculations were done on MareNostrum 4 supercomputer using a single computing node which has 2 sockets Intel Xeon Platinum 8160 CPU with 24 cores each @ 2.10GHz, providing a total of 48 cores. To speed up the calculation of the Transfer entropy the code was parallelized using the Multi- processing Python library. All the code used in this work can be found in the following repository: https://gitlab.bsc.es/flowmaps/te-epidemics.

## Data Availability

No new data was generated as part of this work. All datasets used in this study are available at https://zenodo.org/communities/flow-maps/. Citations for each individual dataset are the following: COVID-19 cases dataset (48); Population dataset (49); Daily Mobility Matrixes dataset (50); and Geographic layer for the different territorial units (51).

## ACKNOWLEDGEMENTS

This work has received funding from the Horizon 2020 project CREXDATA (ID: 101092749) and UNICO I+D Cloud MePreCisa, funded by MINECO and the EU- Next Generation EU, within the framework of the PRTR and the MRR. CP has received funding from the program “Juan de la Cierva - Formación” of the Ministry of Education and Science of Spain (ID: FJC2021-046655-I).

## Notes

### Competing Interest Statement

The authors have declared no competing interest.

