## Supplementary Material for "Unravelling Causal Associations between Population Mobility and COVID-19 Cases in Spain: a Transfer Entropy Analysis"

### Supplementary Information

**Mobility-based TE in epidemiological simulations.** We utilized a metapopulation model known as Susceptible-Infected-Recovered (SIR) model (Fig. S1A), as presented in the work by Tizzoni et al. (13). This model was employed to simulate the progression of an epidemic similar to COVID-19. Our focus was on employing mobility-based Transfer Entropy (TE) to assess how specific subpopulations influence the outbreak in neighboring areas. To achieve this, we conducted simulations using various commuting network structures (Fig. S1B). The aim was to quantify the flow of information between distinct subpopulations during an epidemic that originates within a single subpopulation. Three commuting scenarios were generated, each consisting of populations of equal size ($N = 10,000$ individuals) and a consistent number of commuters ($N_{ij} = 500$) across subpopulations. In all cases, the simulations commenced with 10 infected individuals localized within a specific subpopulation.

We ran epidemic simulations for each mobility network configuration, considering different values for the basic reproductive number ($R_0$): $R_0 = 1.5$, $R_0 = 2.0$, and $R_0 = 3.0$. These values correspond to varying levels of transmissibility, representing a moderately transmissible, highly transmissible, and extremely transmissible epidemic agent, respectively. This range was selected because COVID-19 outbreaks have been associated with $R_0$ values between 2.43 and 3.10 (53). The recovery rate was established as $\mu^{-1} = 3.0$, following the approach outlined by Brockmann (14). Furthermore, we studied the epidemiological dynamics using three different metapopulation structures defined by the mobility networks of daily commuters. We use different mobility network topologies of increasing complexity including a simple chain, a ring, a star and finally a topology based on realistic mobility and population data from Spain (Fig. S1C). The simulations were conducted using the Python Package Epicomute available at https://github.com/franksh/EpiCommute/. Since the simulations are stochastic, for each scenario, we ran 1000 replicates and considered the average trajectory.

We first analysed the characteristic time of the process for each scenario by measuring the total net transfer of entropy using different values for $\omega$ and $\delta$ (Table S1). The results show that characteristic time depends on both the topology of the mobility network as well as the epidemiological parameter $R_0$. Interestingly, we observe that in all the analysed scenarios, the characteristic time of the process defined by the optimal $\omega$ varies in a narrow interval of between 5 and 9 days. On the other hand, the optimal $\delta$ strongly depends on both the topology of the mobility network and the $R_0$. The results showed that the optimal $\delta$ decreases with larger values of $R_0$, as expected. Nevertheless, we did not find any relationship with respect to the mobility network topology. The total net TE tends to increase for more interconnected mobility topologies, i.e. the star-like topology, and to decrease with higher values of $R_0$. Surprisingly, we observe higher net TE when setting a peripheric node (M2) as the epidemic hub instead of the centre node (M1).

**Table S1.** Optimal TE parameters for different SIR simulations.

| Mobility network topology | $R_0$ | Hub | $\delta^*$ | $\omega^*$ | Total $DI^+$ |
|---|---|---|---|---|---|
| chain | 1.5 | M1 | 6 | 6 | 179.9 |
| ring | 1.5 | M1 | 6 | 6 | 193.3 |
| star | 1.5 | M1 | 7 | 8 | 240.4 |
| chain | 2.0 | M1 | 4 | 9 | 157.7 |
| ring | 2.0 | M1 | 5 | 9 | 173.6 |
| star | 2.0 | M1 | 4 | 5 | 220.1 |
| star | 2.0 | M2 | 5 | 8 | 268.7 |
| chain | 3.0 | M1 | 3 | 8 | 92.0 |
| ring | 3.0 | M1 | 3 | 7 | 94.2 |
| star | 3.0 | M1 | 1 | 5 | 96.9 |

Using the optimal parameters that better describe the characteristic time of the epidemic process in each different scenario, we calculated the TE to study the flow of information during the epidemic process. We found that the net transfer of entropy between the subpopulations correctly uncovers the spreading patterns in all topologies analysed.

We first analysed the star-like topology with a peripheral infection hub (Fig. S2A). The epidemic process lasts for around 70 days, with the first wave of infections unfolding in the hub region and then spreading to all other regions through the central node (Fig. S2B). When we calculated the TE over the whole process, we found that the net flow of information $DI^+$ between the subpopulations correctly reconstructed the structure of the spreading process (Fig. S2C). When we perform the same analysis but calculate the transfer of entropy between the incidence time series in different regions, without explicitly taking the mobility patterns into account, non-causal (indirect) correlations can be observed in the entropy flow graph (Fig. S2D). The results considering mobility matrices with chain-like and ring-like topology show the same trends, with the net flow of TE reproducing the topology of the mobility matrix when explicitly taking the mobility into account (Figs. S3 and S4).

DRAFT

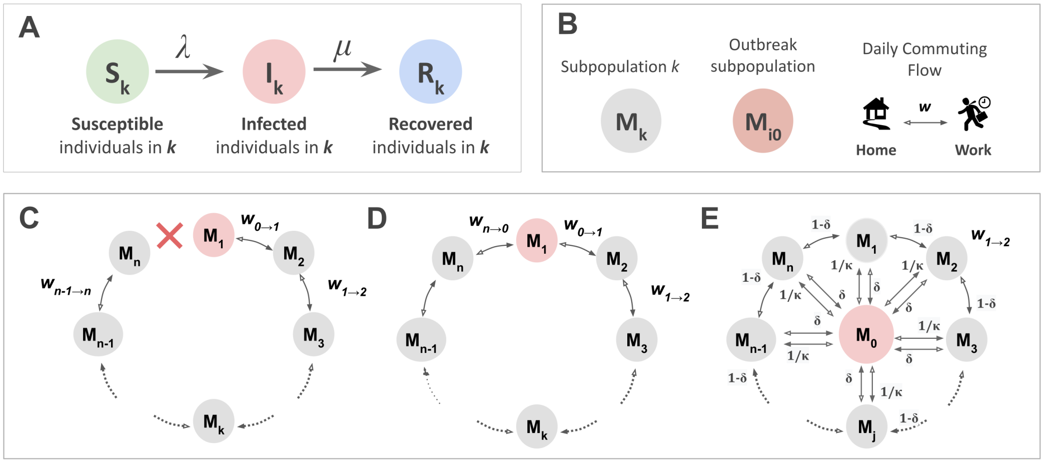


**Fig. S1.** Finding the characteristic time scale of the spreading process of a SIR metapopulation model with different commuting topologies. Panels A, shows the SIR model used to conduct the epidemiological simulations. Panel B shows the basic components of the metapopulation model. including the subpopulations and the commuting flows. Panel C depicts three different topologies for the metapopulation and daily commuting network; from left to right the chain, ring and star topologies.

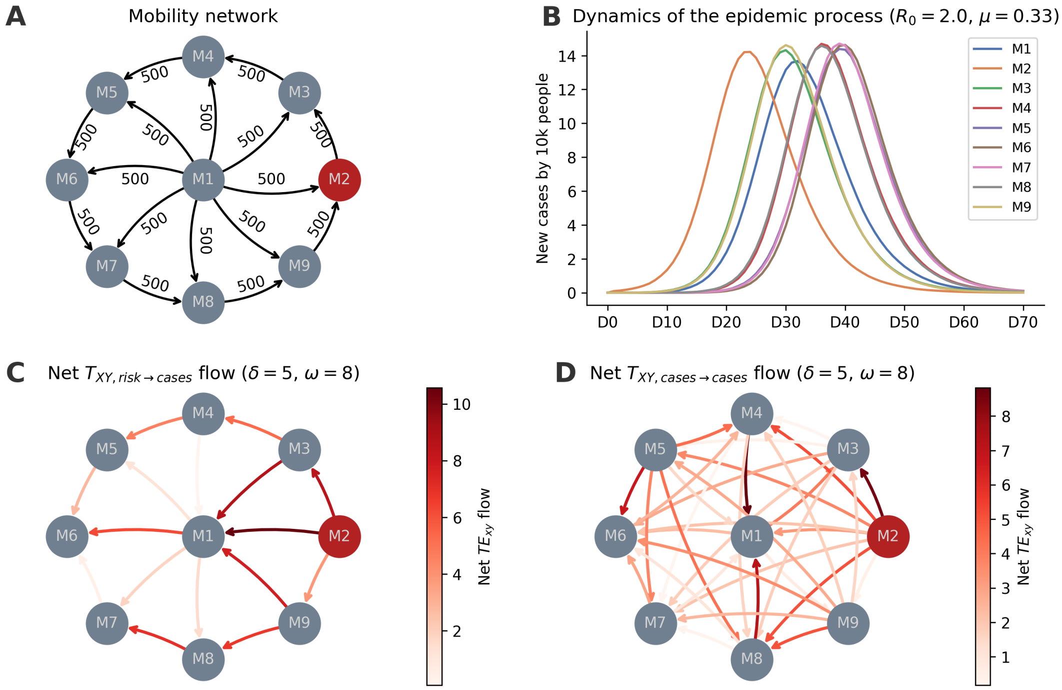


**Fig. S2.** Application of Transfer Entropy to infer a causal relationship between population mobility and the spread of an epidemic in a metapopulation SIR model. Panel A shows the topology of the commuting network, each node has a population of 10k inhabitants, and the number of daily commuters is indicated in the arrows. Panel B shows the average over 1000 simulations of the normalised number of cases as a function of time (in days) obtained using an infection rate $R_0 = 2$ and a recovery rate $\mu^{-1} = 3$. Panels C and D show the net value of $TE_{XY}$ transferred during the entire simulated epidemic for each pair of subpopulations, considering $TE_{risk \to cases}$ and $TE_{cases \to cases}$, respectively. Optimal TE parameters $\delta^* = 5$ and $\omega^* = 8$ were used, the red node is the hub where the epidemic started ($I_0 = 10$).

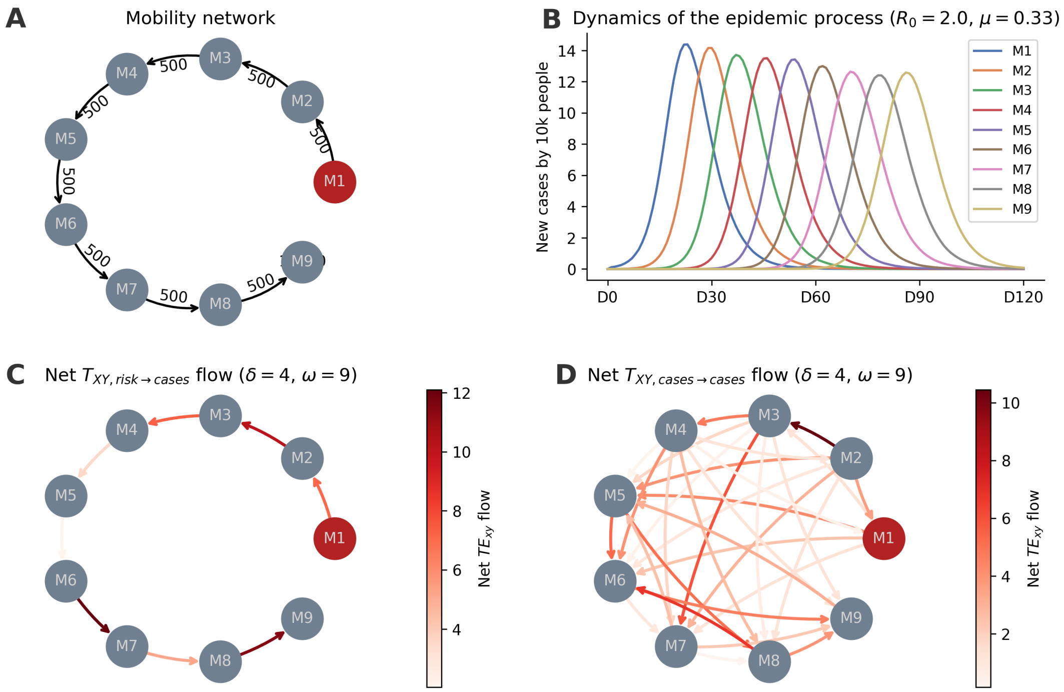


**Fig. S3.** Application of Transfer Entropy to infer a causal relationship between population mobility and the spread of an epidemic in a metapopulation SIR model. Panel A shows the topology of the commuting network, each node has a population of 10k inhabitants, and the number of daily commuters is indicated in the arrows. Panel B shows the average over 1000 simulations of the normalised number of cases as a function of time (in days) obtained using an infection rate $R_0 = 2$ and a recovery rate $\mu^{-1} = 3$. Panels C and D show the net value of $TE_{XY}$ transferred during the entire simulated epidemic for each pair of subpopulations, considering $TE_{risk \to cases}$ and $TE_{cases \to cases}$, respectively. Optimal TE parameters $\delta^* = 4$ and $\omega^* = 9$ were used, the red node is the hub where the epidemic started ($I_0 = 10$).

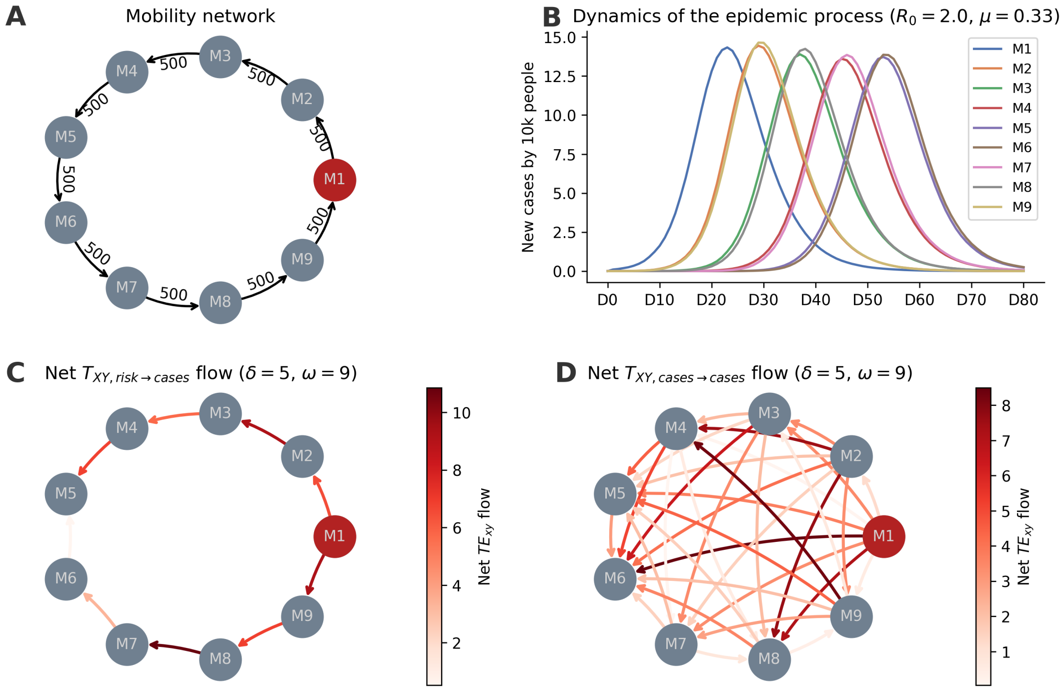


**Fig. S4.** Application of Transfer Entropy to infer a causal relationship between population mobility and the spread of an epidemic in a metapopulation SIR model. Panel A shows the topology of the commuting network, each node has a population of 10k inhabitants, and the number of daily commuters is indicated in the arrows. Panel B shows the average over 1000 simulations of the normalised number of cases as a function of time (in days) obtained using an infection rate $R_0 = 2$ and a recovery rate $\mu^{-1} = 3$. Panels C and D show the net value of $TE_{XY}$ transferred during the entire simulated epidemic for each pair of subpopulations, considering $TE_{risk \to cases}$ and $TE_{cases \to cases}$, respectively. Optimal TE parameters $\delta^* = 5$ and $\omega^* = 9$ were used, the red node is the hub where the epidemic started ($I_0 = 10$).

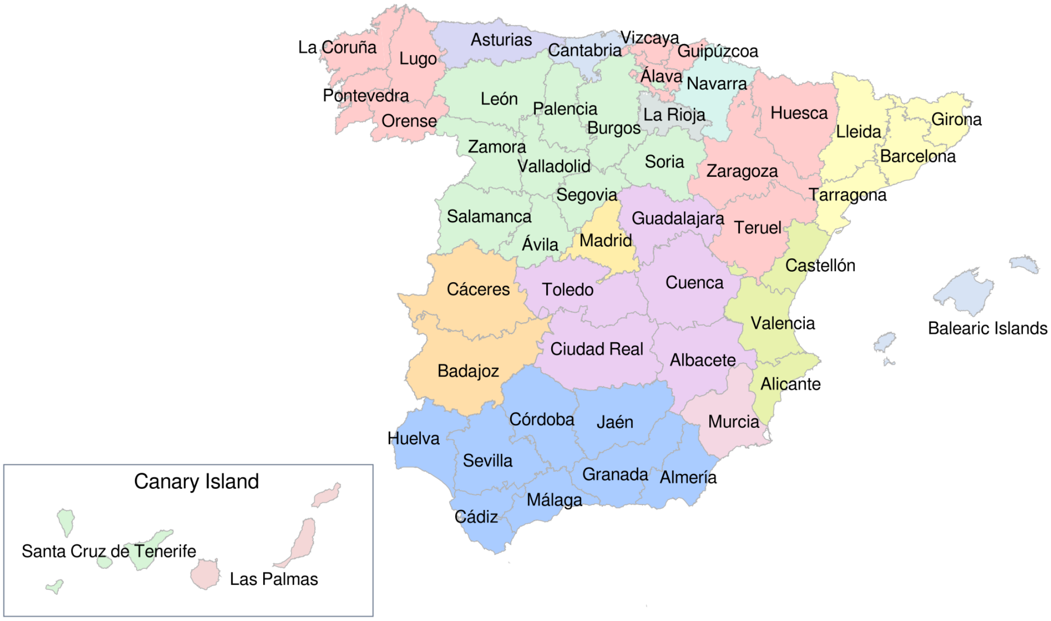


**Fig. S5.** Spanish map of regions and provinces. Each territorial unit corresponds to a province and neighbour provinces of the same color correspond to an Autonomous Community.

DRAFT

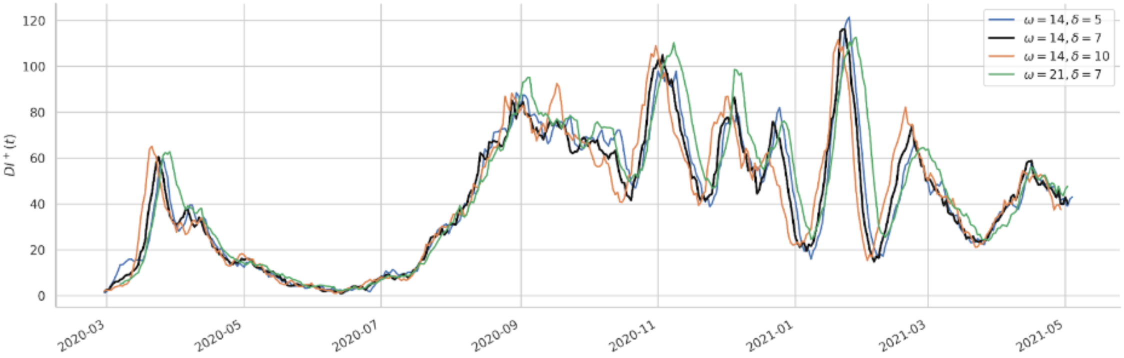


**Fig. S6.** Sensitivity analysis of the $DI^+$ profiles to changes in $\omega$ and $\delta$ parameters. Each curve shows the total $DI^+$ transferred using different values for $\omega$ and $\delta$.

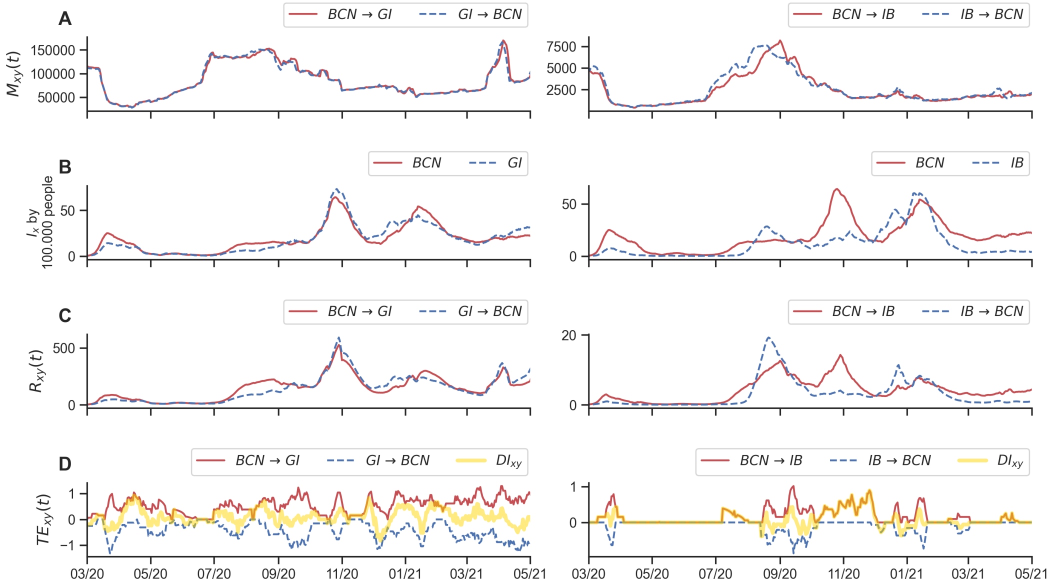


**Fig. S7.** Time series of mobility, cases, and $TE$ between neighbour and non-neighbour provinces. The figure depicts different time series between a pair of neighbour provinces Barcelona-Girona (right panels), and non-neighbour regions Barcelona-Balearic Islands (left panels). From top to bottom, the figure depicts the number of daily trips (A), normalized new cases (B), the associated risk between (C) and the transfer of entropy $TE_{x,y}$ (D). For visualization purposes, $TE_{x,y}$ is represented as positive values whereas $TE_{x,y}$ (the opposite direction) is represented as negative ones. The yellow line in D represents the directionality index $DI_{x,y}$. In the legends, the labels BCN, GI and IB, correspond to Barcelona, Girona and Balearic Islands, respectively.

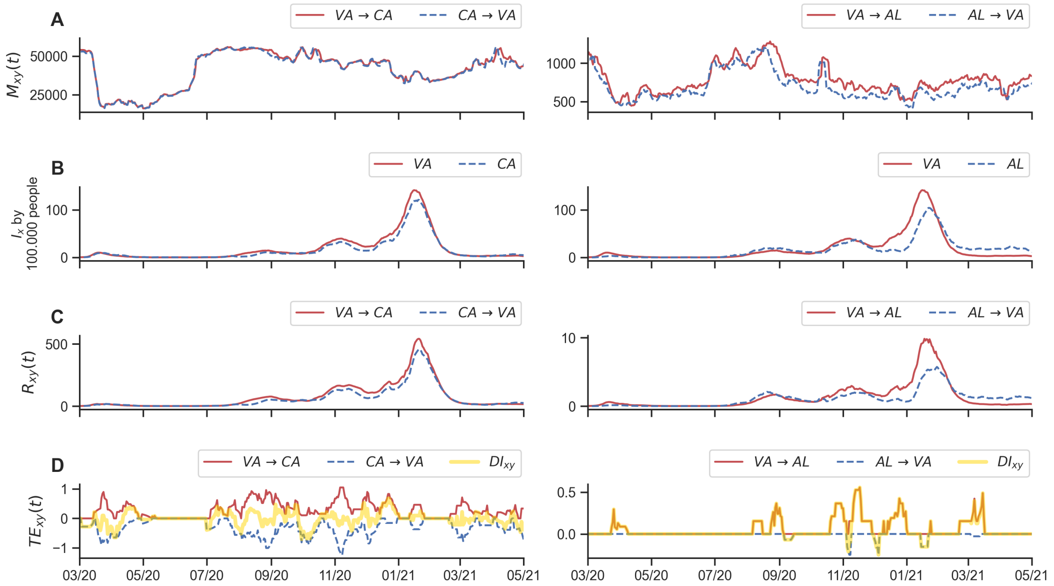


**Fig. S8.** Time series of mobility, cases, and $TE$ between neighbour and non-neighbour provinces. The figure depicts different time series between a pair of neighbour provinces Valencia-Castellón (right panels), and non-neighbour regions Valencia-Almería (left panels). From top to bottom, the figure depicts the number of daily trips (A), normalized new cases (B), the associated risk between (C) and the transfer of entropy $TE_{x,y}$ (D). For visualization purposes, $TE_{x,y}$ is represented as positive values whereas $TE_{x,y}$ (the opposite direction) is represented as negative ones. The yellow line in D represents the directionality index $DI_{x,y}$. In the legends, the labels VA, CA and AL, correspond to Valencia, Castellón and Almería, respectively.

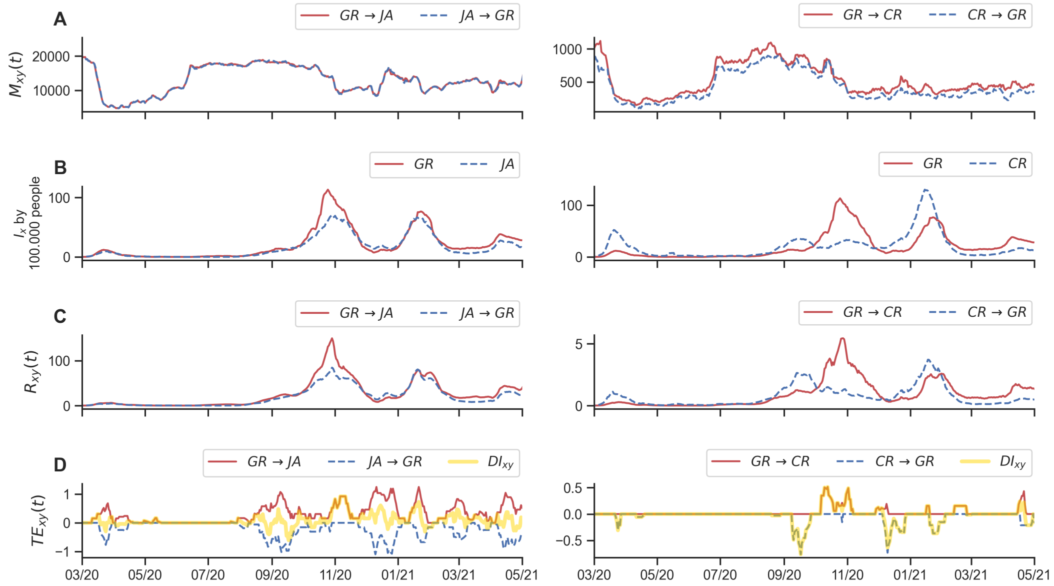


**Fig. S9.** Time series of mobility, cases, and $TE$ between neighbour and non-neighbour provinces. The figure depicts different time series between a pair of neighbour provinces Granada-Jaén (right panels), and non-neighbour regions Granada-CiudadReal (left panels). From top to bottom, the figure depicts the number of daily trips (A), normalized new cases (B), the associated risk between (C) and the transfer of entropy $TE_{x,y}$ (D). For visualization purposes, $TE_{x,y}$ is represented as positive values whereas $TE_{x,y}$ (the opposite direction) is represented as negative ones. The yellow line in D represents the directionality index $DI_{x,y}$. In the legends, the labels GR, JA and CR, correspond to Granada, Jaén and Ciudad Real, respectively.

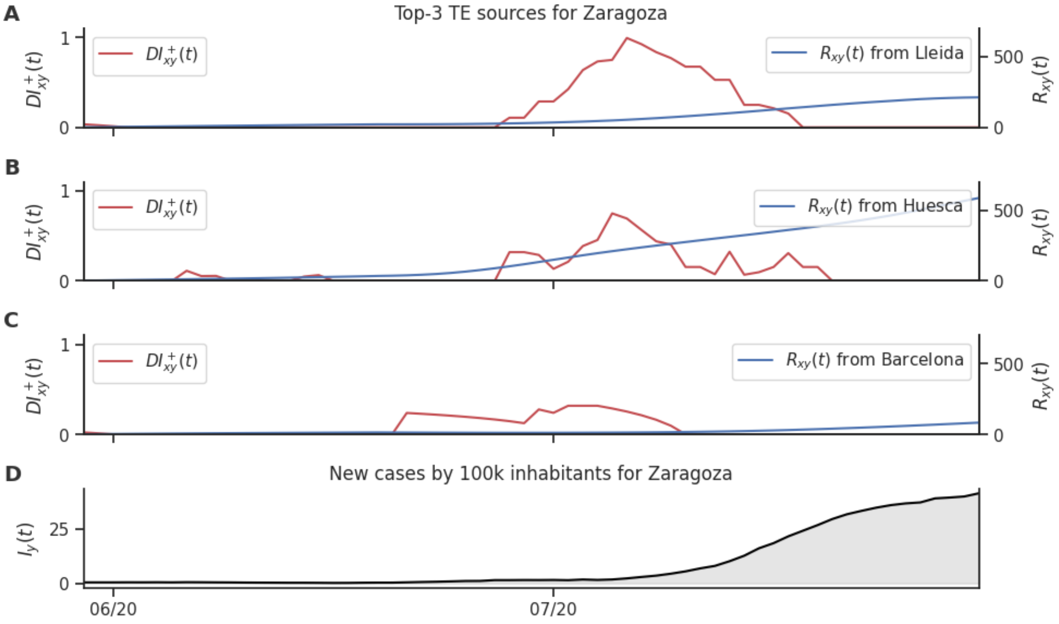


**Fig. S10.** Top DI sources for Zaragoza between June 1st and July 31, 2020. Panels A to C show the net information transfer $DI^+_{xy}(t)$ (in red) and the mobility-realted risk $R_{xy}(t)$ (in blue) for each one of Zaragoza's top DI sources during the months of June and July. Panel D shows the number of new COVID-19 cases in Zaragoza during the same period.

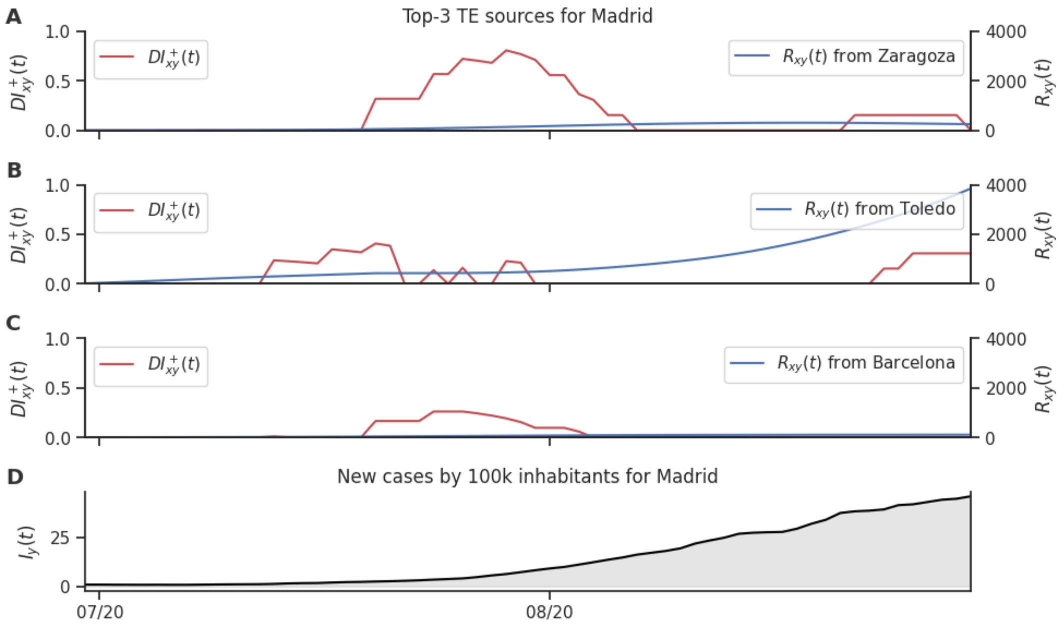


**Fig. S11.** Top DI sources for Madrid between July 1st and August 31, 2020. Panels A to C show the net information transfer $DI^+_{xy}(t)$ (in red) and the mobility-realted risk $R_{xy}(t)$ (in blue) for each one of Madrid's top DI sources during the months of July and August. Panel D shows the number of new COVID-19 cases in Madrid during the same period.

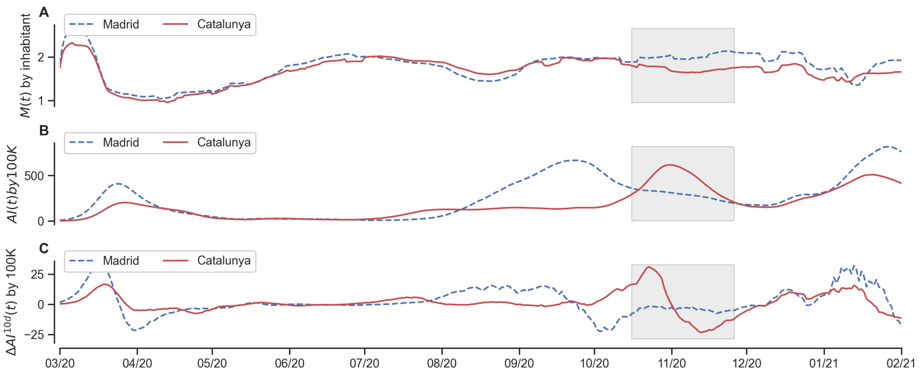


**Fig. S12.** Time series of cases and mobility for Catalunya and Madrid. Panel A shows the number of trips $M(t)$ per inhabitant. Panel B shows the accumulated incidence $AI^{10}$ over the last ten days by 100,000 inhabitants. Panel C shows the rate of change of $AI^{10}$, calculated using a first-order finite difference approximation. The grey rectangle indicates the period in which the policy of closing bars and restaurants was applied in Catalunya; the period of the application of the policy spanned between 2020-10-16 and 2020-11-26.

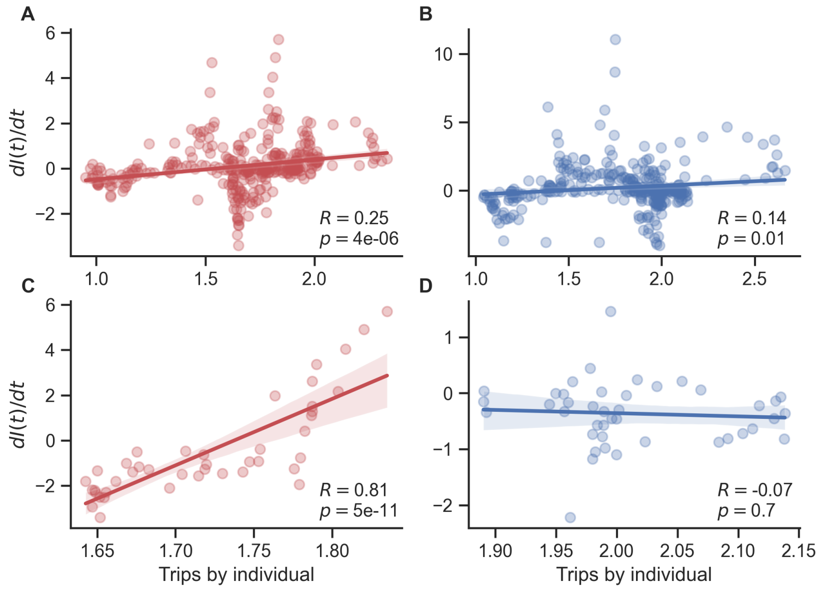


**Fig. S13.** Correlations between the number of trips $M(t)$ per inhabitant and the rate of change of $AI^{10}$. Panels A and B show the relations between the number of trips $M(t)$ per inhabitant and the rate of change of $AI^{10}$ for Catalunya and Madrid respectively considering a period of time between March 2020-03-01 and 2021-02-01. Panels C and D show the relations between the number of trips $M(t)$ per inhabitant and the rate of change of $AI^{10}$ for Catalunya and Madrid respectively, considering the period in which the policy of closing bars and restaurants was applied in Catalunya; the period of the application of the policy spanned between 2020-10-16 and 2020-11-26. The rate of change of $AI^{10}$ was calculated using a first-order finite difference approximation.

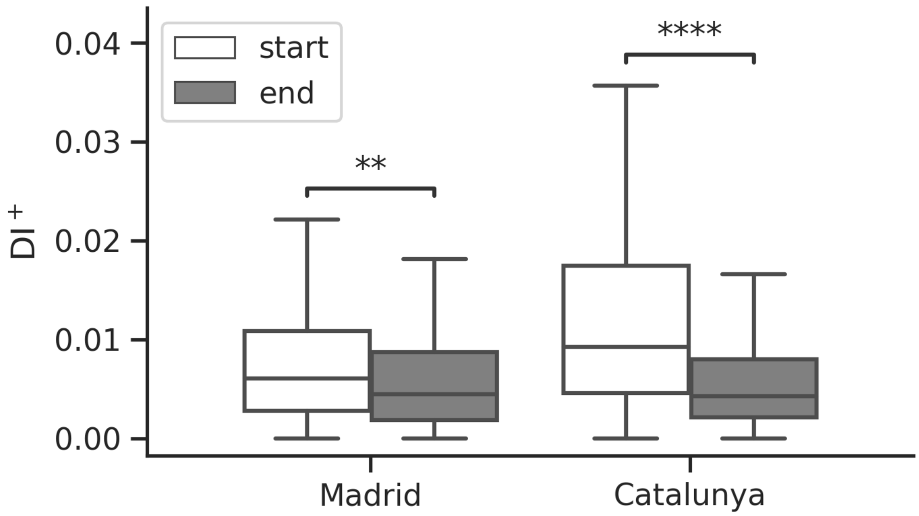


**Fig. S14.** Impact of the temporary closing of bars and restaurants in Catalonia. Difference in the distribution of average DI+ per BHA values in the 2 weeks following the start of the closing of bars policy (start), and in the last 2 weeks of the policy (end). Comparison between the autonomous communities of Madrid (where no policy was applied, p = 4.7e-03, Mann-Whitney test) and Catalonia (where the policy was applied, p = 2.3e-22, Mann-Whitney test).

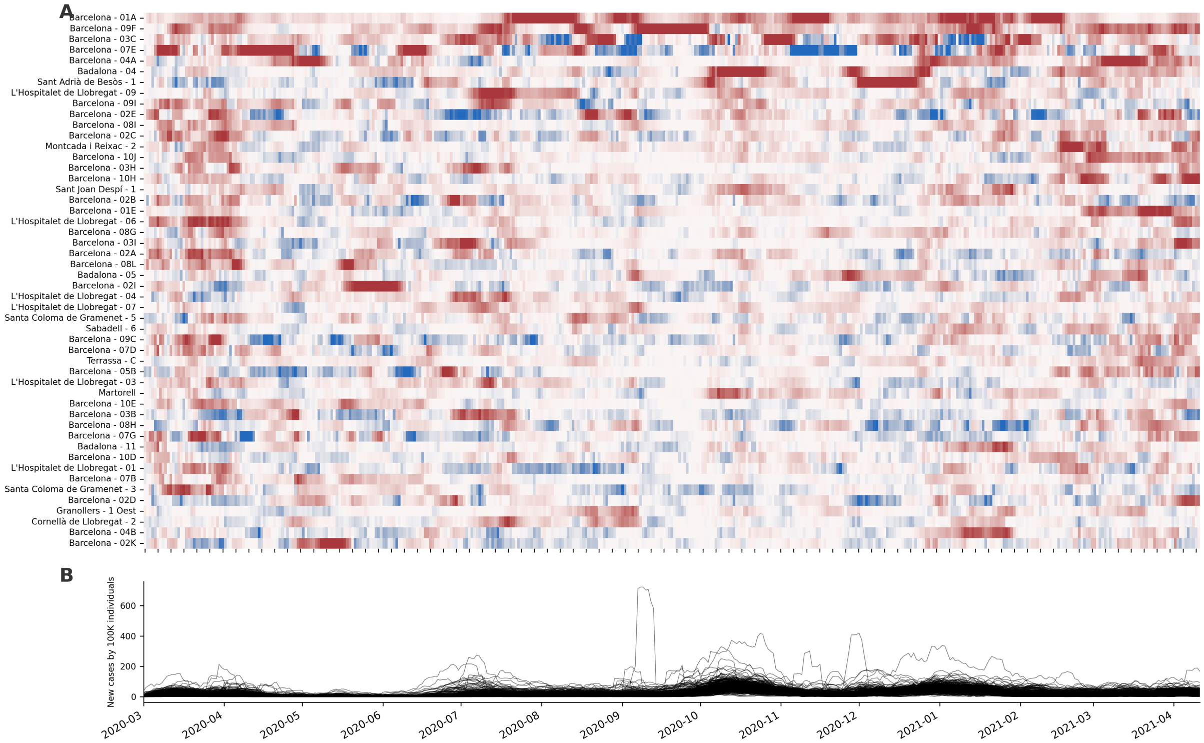


**Fig. S15.** Dynamics of the COVID-19 pandemic in Catalunya. Panel A shows the normalised directionality index ($DI_x$) for basic health areas in Catalunya calculated using parameters $\delta = 7$ days and $\omega = 14$ days. Provinces are ordered by total $DI_x$; only the top-50 drivers are shown. Panel B shows the average number of daily COVID-19 by 100k inhabitants for the same period.
